# Occipital gamma-aminobutyric acid and glutamate-glutamine alterations in major depressive disorder: An MRS study and meta-analysis

**DOI:** 10.1101/2020.11.26.20239491

**Authors:** Vuong Truong, Paul Z. Cheng, Hsin-Chien Lee, Timothy J. Lane, Tzu-Yu Hsu, Niall W. Duncan

## Abstract

The neurotransmitters GABA and glutamate have been suggested to play a role in Major Depressive Disorder (MDD) through an imbalance between cortical inhibition and excitation. This effect has been highlighted in higher brain areas, such as the prefrontal cortex, but has also been posited in basic sensory cortices. Based on this, magnetic resonance spectroscopy (MRS) was used to investigate potential changes to GABA+ and glutamate+glutamine (Glx) concentrations within the occipital cortex in MDD patients (n = 25) and healthy controls (n = 25). No difference in occipital GABA+ or Glx concentrations, nor in the GABA+/Glx ratio, was found between groups. An analysis of an extended MDD patient and unmatched control dataset (n = 90) found no correlation between metabolite concentrations and depressive symptoms. These results were integrated with prior studies through metabolite-specific meta-analyses, revealing no difference in occipital GABA and Glx concentrations between patients and controls. An effect of publication year on GABA group differences was found, suggesting that previously reported results may have been artifacts of measurement accuracy. Taken together, our results suggest that, contrary to some prior reports, MRS measurements of occipital GABA and Glx do not differ between MDD patients and controls.

## 1. Introduction

Major depressive disorder (MDD) is a serious psychiatric disorder that is highly prevalent worldwide and contributes significantly to the global disease burden (James et al., 2018). Although there has been extensive research into MDD over many decades, concrete biological mechanisms behind this heterogeneous disorder have not been established (Belmaker and Agam, 2008). Traditionally, a large proportion of MDD research has focused on a monoamine-deficiency model of depression, with mixed success (Boku et al., 2018; Hirschfeld, 2000). In more recent years this has been joined by a number of alternative models including, for example, ones that focus on stress and glucocorticoids (Hirschfeld, 2000), neurotrophic factors (Price and Duman, 2020), or inflammation (Woelfer et al., 2019).

Another influential hypothesis proposes changes to the glutamatergic and GABA-ergic systems in MDD. These represent, respectively, the main excitatory and inhibitory transmitters in the human brain. Focusing on GABA, direct observations have suggested that its concentration is reduced in the plasma, cerebrospinal fluid, and brain tissue samples of MDD patients (Croarkin et al., 2011). This picture is less clear, however, when non-invasive magnetic resonance spectroscopy (MRS) has been used to measure GABA concentrations in MDD patients in vivo. Although some such studies have also found GABAergic reductions, others have reported no such effect (Godfrey et al., 2018). MRS studies of glutamate concentrations in MDD have reported similarly conflicting results (Godfrey et al., 2018).

These inconsistencies in reported differences between patients and controls may arise from the apparent independence of metabolite concentrations across different brain regions within individuals (Duncan et al., 2019; Greenhouse et al., 2016). This means that an alteration in one region, or in amalgamated whole-brain measures, may not necessarily indicate an alteration in any other specific region. A further cause of inconsistent findings may be the range of MRS sequences that have been used across prior studies (e.g., SPECIAL, J-Editing, or PRESS) as these have variable sensitivity to different metabolites. Given these issues, there is a need to replicate results for consistent regions of interest (ROIs) using appropriate MRS techniques, as well as a need to combine existing results through meta-analyses in order to resolve inconsistencies.

Previously, a large proportion of MDD research focused on “higher” brain areas such as the anterior cingulate cortex (ACC) and prefrontal cortex due to their involvement in processes, such as emotion regulation, that are commonly implicated in MDD pathophysiology (Park et al., 2019). More recently, attention has also switched to investigate primary sensory cortices, in part in response to alternative theories, such as the so-called “predictive coding” or “active inference” accounts of psychopathology that incorporate a role for early sensory processing in the disease process (Barrett et al., 2016; Kube et al., 2020). Within this framework, the primary visual cortex plays an important role in processing information related to emotion (Barrett and Bar, 2009; Kragel et al., 2019) and other higher cognitive processes such as attention, working memory, and decision making (Roelfsema and de Lange, 2016). A number of studies have used MRS to investigate GABA and glutamate within the occipital cortex in MDD. Results have, however, been inconsistent, with some studies finding metabolite reductions (Bhagwagar et al., 2007; Sanacora et al., 2004, 1999) but others finding no difference between patients and controls (Price et al., 2009; Shaw et al., 2013). This inconsistency points to a need for additional studies targeting this region, along with a synthesis of existing results through a meta-analysis of occipital cortex MRS studies in MDD.

Based on this, we acquired MRS data from the occipital cortex in a group of MDD patients to compare GABA and glutamate+glutamine (Glx) concentrations against a group of age and sex matched controls. The ratio between GABA and Glx was also compared. These results were then integrated with those of prior studies investigating this region in MDD through separate meta-analyses of GABA and Glx group differences. In addition to these group comparisons, we also conducted a parametric analysis of depressive symptoms against metabolite concentrations in a larger sample consisting of additional MDD patients and unmatched control participants. This supplementary analysis investigated how GABA and Glx concentrations may be related to symptom load without imposing a categorical classification.

## 2. Experimental procedures

### 2.1 Participants

Twenty-five patients diagnosed with MDD (20 females, age 36.6 ± 11.9 years old) were recruited from the out-patient department of the Department of Psychiatry, TMU-ShuangHo Hospital, Taiwan. Their current depressive episode was confirmed using the Mini International Neuropsychiatric Interview (Sheehan et al., 1998). All patients were undergoing pharmacotherapy at the time of scanning. Time since onset of depressive episodes and medications varied across patients (see Table S1 for details). Overall, the patients had an average time since their first depressive episode onset of 10.3 ± 11.1 years. The average duration of their current depressive episode was 9.1 ± 16.5 months. Patients prescribed GABAergic hypnotics were asked to not use them the night before scanning. Twenty-five healthy control participants (20 females, age 37.0 ± 11.5 years old) with no history of psychiatric disorders were recruited from the local community. This group were directly matched for age and sex with MDD patients. The main analyses of this paper used this matched MDD-control dataset.

To further investigate the relationship between depressive symptoms and metabolite concentrations, additional MDD and control participants were recruited according to the same inclusion criteria as the matched group. These additional participants were not matched for age and sex. The extended dataset thus includes the previously described matched MDD-control participants plus newly-recruited participants to give a total of 33 MDD patients (25 females, age 38.8 ± 13.3 years old) and 62 healthy controls (51 females, age 32.9 ± 11.0 years old).

Exclusion criteria for all participants were: contraindications for magnetic resonance imaging (e.g., brain implants, cardiac pacemakers); pregnancy; a history of neurological disorders (e.g., stroke, seizure, traumatic brain injury); comorbid psychiatric conditions; and alcohol or substance dependence. All participants received both oral and written information regarding the purpose and procedures of the study. This information was given in quiet surroundings and participants had the opportunity to ask questions about their participation in the study. Written consent was obtained following this. The study was approved by the Taipei Medical University Joint Institutional Review Board (N201603080).

### 2.2 Symptom measures

Patient symptom severity on the day of scanning was assessed with the MADRS instrument (Montgomery and Asberg, 1979). Depressive symptoms across patients and controls were measured using the Beck Depression Inventory-II (BDI) (Beck et al., 1996).

### 2.3 MRI acquisition

MRS data were acquired on a GE MR750 3-Tesla system using a body-coil for transmission and an 8-channel receive head-coil. A high resolution T1-weighted 3D structural Image (FSPGR; spatial resolution = 1 × 1 x 1 mm^3^; FoV = 256 × 256 mm^2^) was firstly acquired. Using this image, an MRS voxel was located in the occipital cortex, with a target volume of 27 mm^3^. The exact size and location of the MRS voxel was adjusted to accommodate individual differences in brain size and morphology whilst avoiding areas with poor signal. Voxel locations are shown in Figure 1A. GABA sensitive MEGA-PRESS data were then acquired using the following settings: TR = 2000 ms; TE = 68 ms; data points = 4096; spectral width = 2000 kHz; alternating ON/OFF editing; 14 ms editing pulses applied at 1.9 ppm (ON) and 7.46 ppm (OFF); 192 averages; 8 water unsuppressed acquisitions. These settings do not suppress the contribution of macromolecules to the signal and so quantified measurements are referred to as GABA+.

**Figure 1:**
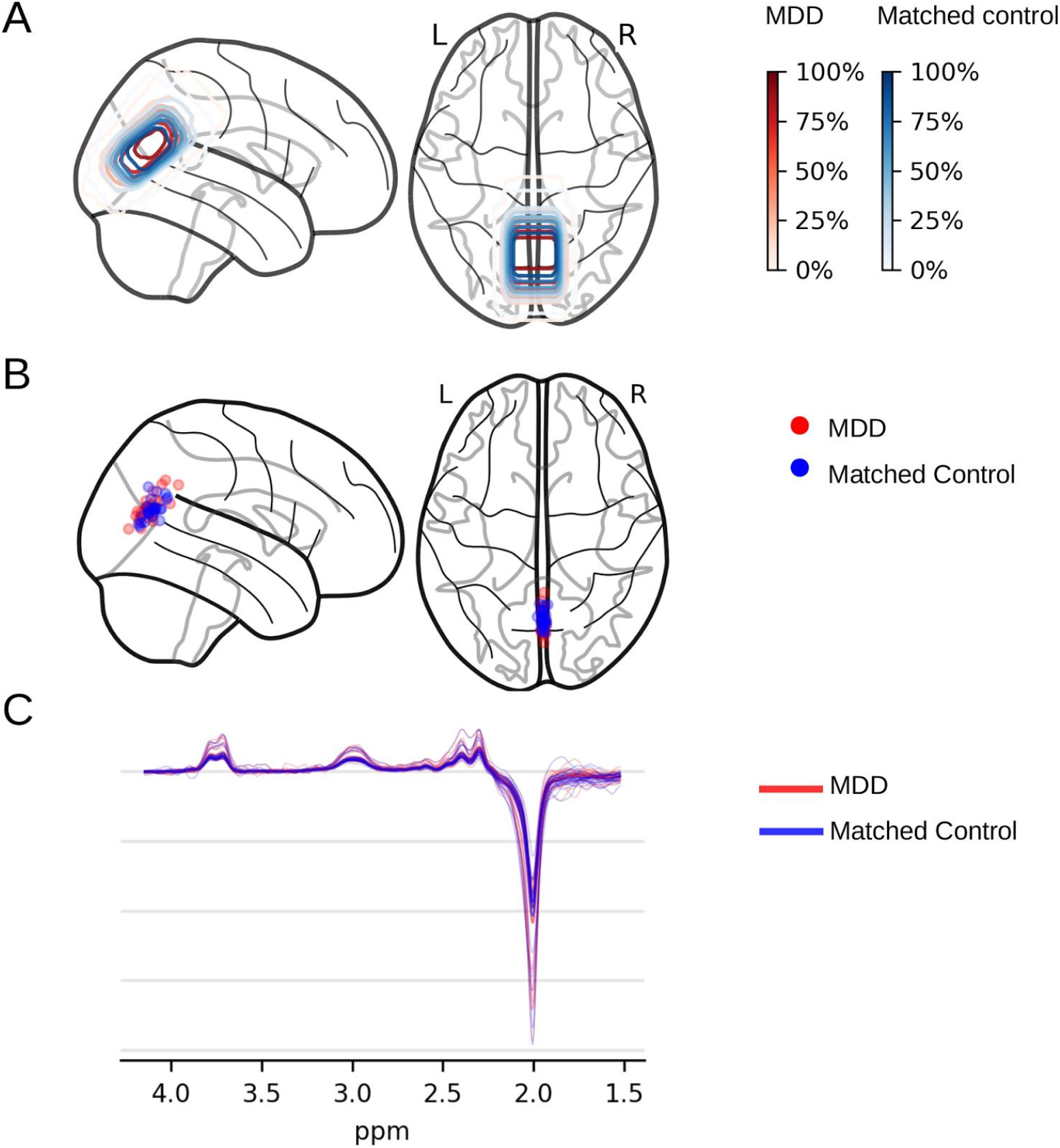
MRS voxel location and MEGA-PRESS difference spectra (Matched MDD–Control dataset). (A) Voxel overlap densities for MDD patients (n = 25) and their matched controls (n = 25), as denoted by red and blue contours. (B) Voxel centroids for each MDD patient (n = 25) and their matched controls (n = 25), as denoted by red and blue colours. Brain outlines were created using the nilearn toolbox (Abraham et al., 2014). (C) Individual MEGA-PRESS difference spectra along with the average spectra for MDD patients (n = 25) and their matched controls (n = 25), as denoted by red and blue colours. Thicker lines represent the group means.

### 2.4 MRI analysis

All spectra were processed using the Gannet 3.0 toolbox (gabamrs.blogspot.co.uk), running in MATLAB R2019a. In brief, the preprocessing of MRS data was done automatically through Gannet’s standard protocol, including frequency and phase correction, fast Fourier transform of time-domain acquired data to frequency-domain spectra, and exponential line broadening. After that, the GABA+ peak at 3.0 ppm in the difference-edited spectrum was fitted with a single Gaussian model and a composite measure of glutamate and glutamine (Glx) was obtained by fitting the Glx peak at 3.7 ppm with a Gaussian doublet model, as described elsewhere (Edden et al., 2013).

MRS voxel coregistration to each subject’s structural image was performed by Gannet, along with tissue segmentation to gray matter (GM), white matter (WM) and cerebrospinal fluid (CSF) probabilistic partial volume maps by the unified tissue segmentation algorithm in SPM12 (fil.ion.ucl.ac.uk/spm). These maps were used to correct for the visibility and relaxation of water signals in GM/WM/CSF and for the neurotransmitter concentration difference between GM and WM. All neurotransmitter concentrations were quantified in institutional units (i.u.) with the water signal as the reference. T1 structural images and MRS voxel masks from all subjects were co-registered to MNI152 standard space using FMRIB Software Library 5.0.11 Linear Image Registration Tool (FSL FLIRT; fsl.fmrib.ox.ac.uk).

### 2.5 Data quality and voxel overlap

For comparison of MRS data between individuals it is important that the voxels from which data are acquired cover the same anatomical locations. Similarly, group comparisons require there to be no systematic difference in the anatomical locations covered for groups (Truong and Duncan, 2020). To identify potential outlying voxel locations we calculated individual voxel centres-of-mass in MNI space and calculated the Mahalanobis distance for each against the group 3D distribution (Martos et al., 2013). Two participants were excluded based on a chi-square probability density function with three degrees of freedom. To ensure there was no group difference (MDD vs controls) in voxel location, we ran a k-means clustering analysis with two clusters on the 3D voxel coordinates and compared this to the diagnosis ground-truth. Group membership could not be estimated based on voxel location, suggesting no overall location difference between groups. MRS spectra were visually appraised to identify poor quality data (Figure 1C). No data were excluded following this. Frequency drift was then calculated and one participant excluded based on a cut-off value of 15.5Hz (Tsai et al., 2016).

### 2.6 Statistical analysis

All statistical analyses were conducted using the Pingouin package (pingouin-stats.org) in Python 3.6 and the WRS2 package (cran.r-project.org/web/packages/WRS2/index.html) in R (version 3.6.1).

MRS estimates of neurotransmitter concentrations (GABA+, Glx, GABA+/Glx ratio) were compared between patients and matched controls by Yuen’s test for trimmed means (Wilcox and Rousselet, 2018). The correlation between metabolite concentrations (GABA+, Glx or GABA+/Glx ratio) and depressive symptoms was then tested in the extended dataset using a robust “percentage bending” method (Wilcox and Rousselet, 2018). This was done with MADRS scores in the extended MDD group (n = 29) and with BDI scores across all participants (n = 90). Participant age was included as a covariate in these correlations.

A number of supplementary analyses were conducted. Firstly, age and sex effects on the GABAergic system have been proposed (Pandya et al., 2019) and so GABA+ and Glx levels were compared between male and female participants and correlated with age in the total extended dataset. Secondly, as some studies have suggested that psychotropic drugs, such as SSRIs or BDZs, might modulate GABA concentrations in the occipital cortex (Bhagwagar et al., 2004; Goddard et al., 2004), we also used a robust ANOVA based on trimmed means to compare GABA concentrations between matched controls, MDD without SSRI or BDZ treatment, and MDD with only SSRI or BDZ treatment. Post-hoc t-tests based on trimmed means were computed where a significant main effect was found.

Welch’s corrections were applied for unequal variances between groups and multiple comparisons were FDR corrected with the Benjamini, Hochberg, and Yekutiel method (Benjamini and Yekutieli, 2001). Descriptive statistics are presented as mean ± standard deviation unless otherwise specified. Effect sizes for t-tests, robust tests on trimmed means, and robust ANOVAs were reported as Cohen’s d, robust-d, and ξ.

### 2.7 Meta-analysis - literature search

Pubmed and Google Scholar databases were searched for publications up to December 2019 using the search terms: (“gamma-aminobutyric acid” or “γ-aminobutyric acid” or “GABA”) and (“magnetic resonance spectroscopy” or “MRS”) and (“major depressive disorder” or “MDD” or “unipolar depression”). Additional publications were identified from the reference lists of two previous meta-analyses (Godfrey et al., 2018; Romeo et al., 2018) that include MRS estimates of occipital GABA or Glx concentrations.

MRS studies considered for inclusion had to be written in English and contain the following information: (1) GABA and/or Glutamate/Glutamine (Glx) concentrations (quantified by MRS) in the occipital cortex of MDD patients; (2) MDD patients had to be diagnosed with an appropriate diagnostic criteria such as Diagnostic and Statistical Manual of Mental Disorders (DSM), MINI, or International Statistical Classification of Diseases and Related Health Problems (ICD); (3) the MRS data in the relevant region was also available in a healthy control group. The inclusion criteria had no restriction on age, gender, mental state, or treatment level. The exclusion criteria included: studies with only a patient or a healthy group; studies which only compared MDD groups with other psychiatric disorders, such as bipolar disorder, schizophrenia, and obsessive-compulsive disorder; or studies with depressive disorders other than MDD (e.g., peripartum depression). This study selection process is summarised in Figure S1.

Information extracted from the selected studies included: Types of metabolites (GABA, glutamate, glutamine, or Glx); concentrations (mean and standard deviation or standard error of the mean); test statistics for group comparisons (if metabolite concentrations were not available); voxel placement location (Occipital), voxel size; number of patients/controls; diagnostic criteria; treatment status at time of scanning; magnetic field strength (Tesla); MRS sequence used; and unit of measurement (mM, mmol/L, metabolite/Cr, metabolite/water, mmol/kg, institutional unit, arbitrary unit). For studies that only reported the concentrations of metabolites as figures, a semi-automated tool (WebPlotDigitizer Version 4.2; automeris.io/WebPlotDigitizer) was utilized to extract the numerical data regarding mean and standard deviation.

Glutamate and glutamine values were summed to give a Glx value in studies that reported these separately. The Glx standard deviation was calculated as the average standard deviation of both glutamate and glutamine (Godfrey et al., 2018).

### 2.8 Meta-analysis - statistical analysis

Data analysis was carried out using the “meta” package in R version 3.6.1. Standardized mean differences (SMDs) with Hedge’s correction (Hedge’s g) between patients and controls were calculated for GABA and Glx concentrations. Hedge’s g and its standard error were calculated using the “escal” R package.

After data preparation, a random-effects model was fitted to the data due to the presence of different magnetic field strengths, MRS sequences, and treatment regimes across studies. The corresponding pooled effect size and its 95% confidence intervals (CI) were calculated from the random-effect model with a Sidik-Jonkman estimator for tau^2^ and Hartung-Knapp adjustment (Röver et al., 2015). Heterogeneity was assessed with Q statistics and was considered significant when I^2^ > 50% or P < 0.1 (Higgins et al., 2003). An Influence Analysis was also conducted to identify any studies that had an extreme impact on the meta-analysis result (as in Supplementary figure S3). The random-effects model was then refitted after excluding any such study. Outliers were identified based on two criteria: (1) having an extreme impact in the Influence Analysis; and (2) having a 95% CI lying outside the pooled effect size 95% CI. A permutation test was used to test the significance of the pooled Hedge’s g as this can reduce the type I error rate compared to the standard Wald and likelihood ratio tests (Follmann and Proschan, 1999). Finally, potential publication bias was assessed by visual inspection of Begg’s funnel plots. A random effects meta-regression analysis was also performed to assess the influence of the publication year and magnetic field strength on the effect sizes found in individual studies.

## 3. Results

### 3.1 MRS data quality

MRS voxel location and spectral quality were compared between match MDD and control groups (see Figure 1 and Table 1). Voxel volumes in the MDD group were larger than controls (*T*_(48)_ = 2.13, *p* = 0.04, *d* = 0.60) but frequency drift did not differ between groups (*T*_(48)_ = 1.43, *p* = 0.16, *d* = 0.40). GABA+ with water fit error did not differ between MDD and control (*T*_(48)_ = −1.60, *p* = 0.12, *d* = 0.45). Similarly, Glx with water fit error did not differ between MDD and control (*T*_(48)_ = −1.74, *p* = 0.09, *d* = 0.49). These fit error values were comparable with other studies using the same MRS sequence (Edden et al., 2013; Mikkelsen et al., 2019).

**Table 1:** MRS data metrics. Details of MRS voxels and MRS spectral quality metrics for matched MDD and control groups.

|  | **MDD**<br>(n = 25) | **Control**<br>(n = 25) |  |
| --- | --- | --- | --- |
| Voxel volume (ml) | 24.4 ± 0.9 | 23.8 ± 1.2 | $T_{(48)} = 2.13, p = 0.04$ |
| Frequency drift (Hz) | 5.2 ± 1.4 | 4.6 ± 1.4 | $T_{(48)} = 1.43, p = 0.16$ |
| GABA fit error (%) | 3.6 ± 1.6 | 4.4 ± 2.0 | $T_{(48)} = -1.60, p = 0.12$ |
| Glx fit error (%) | 3.0 ± 0.9 | 3.5 ± 1.3 | $T_{(48)} = -1.74, p = 0.09$ |

### 3.2 Group differences in occipital cortex GABA+ and Glx

Estimated GABA+ and Glx concentrations were compared between patients (n = 25) and controls (n = 25) from the matched dataset (Figure 2). No differences were found for these (GABA+: *T*_(26.77)_ = 0.54, *p*_FDR_ = 0.89, d = 0.17; Glx: *T*_(27.91)_ = 0.12, *p*_FDR_ = 0.90, *d* = 0.04), nor for the GABA+/Glx ratio (*T*_(26.52)_= 0.68, *p*_FDR_ = 0.89, d = 0.21).

**Figure 2:**
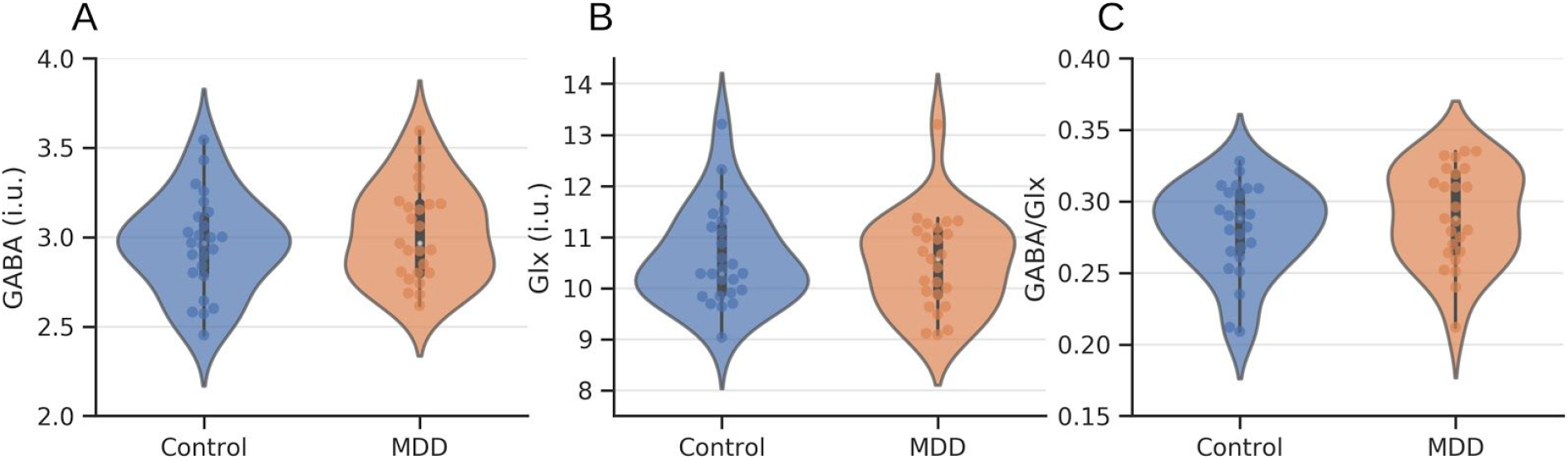
MDD and control metabolite differences. No differences were found between matched patients (n = 25) and controls (n = 25) for estimates of (A) GABA+, (B) Glx, and (C) GABA+/Glx ratios. MDD patients are shown in orange and controls in blue. Box plots indicate medians and quartile ranges. Concentrations are given in institutional units (i.u.).

### 3.3 Factors influencing GABA+ and Glx estimates

The extended dataset of unmatched MDD (n = 29) and control (n = 61) participants was used to investigate factors that correlate with GABA+ and Glx concentration estimates. No influence of sex was found on either GABA+ (*T*_(16.03)_ = 1.30, *p*_FDR_ = 0.42, *d* = 0.34) or Glx (*T*_(21.11)_ = 0.80, *p*_FDR_ = 0.43, *d* = 0.18; Figure 3A). Age was found to correlate with GABA+ estimates (*r* = 0.25, *p*_FDR_ = 0.03; Figure 3B), but not with Glx (*r* = 0.08, *p*_FDR_ = 0.47). No relationship was found across all participants between depressive symptoms, as measured with the BDI scale, and metabolite estimates while controlling for age (GABA+: *r* = 0.11, *p*_FDR_ = 0.58; Glx: *r* = 0.07, *p*_FDR_ = 0.58; GABA+/Glx: *r* = 0.06, *p*_FDR_ = 0.58). In MDD patients, no significant correlation between MADRS scores and GABA+ (*r* = −0.23, *p*_FDR_ = 0.26), Glx (*r* = 0.22, *p*_FDR_ = 0.26), or GABA+/Glx ratios (*r* = −0.36, *p*_FDR_ = 0.20) was found after controlling for age.

**Figure 3:**
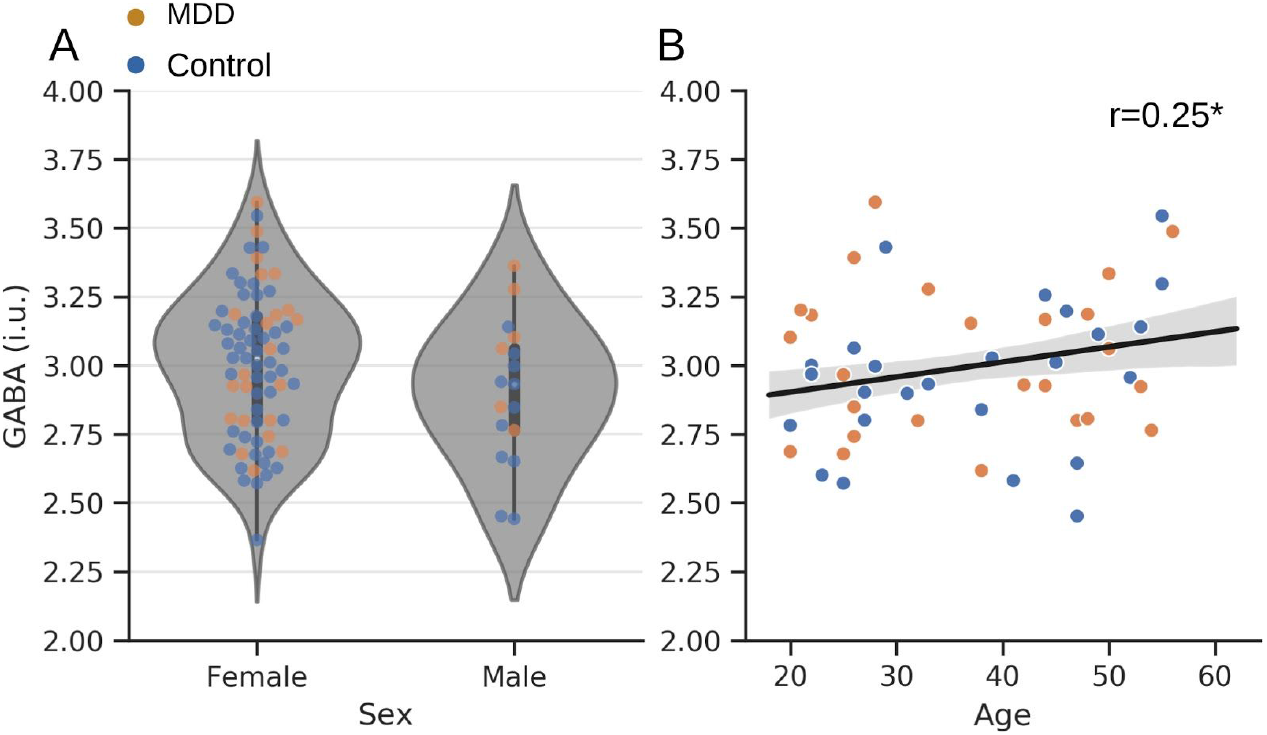
Relation of occipital GABA+ to age and sex. (A) GABA+ concentrations were compared between female (n = 73) and male (n = 17) participants. No difference was observed. (B) A positive correlation between participant age and GABA+ concentrations was found (n = 90). MDD patients are shown in orange and controls in blue. The best linear fit to the data is shown with its 95% CI. Box plots indicate medians and interquartile range. * denotes *p*_FDR_ < 0.05.

Testing the influence of medication on metabolite estimates, a robust ANOVA showed that there was no difference in the occipital GABA+ concentrations across healthy controls, MDD patients undergoing SSRI treatment, and MDD patients not undergoing SSRI treatment (*F*_(2,12.13)_ = 1.38, *p* = 0.29, ξ = 0.42; Supplementary figure S4A). Similarly, occipital GABA+ concentrations did not differ across healthy controls, MDD patients undergoing BDZ treatment, and MDD patients not undergoing BDZ treatment (*F*_(2,14.33)_ = 0.16, *p* = 0.85, ξ = 0.12; Supplementary figure S4B).

### 3.4 Meta-analysis

The literature search identified a total of 193 potential studies (see Figure S1 for an overview). An initial abstract screening reduced this to 17 studies, from which a further six were removed according to the defined exclusion criteria (details of excluded studies are given in Table S2). A total of 12 studies (eleven previous studies, plus this work) were thus included in the meta-analyses (Table 2). Nine of these reported GABA results and eight reported Glx (some reporting both). One outlying GABA study (Sanacora et al., 1999) was identified based on study heterogeneity and excluded from the meta-analysis (before exclusion *I*^2^ = 70.5%, *p* < 0.001; after exclusion, *I*^2^ = 25.8%, *p* = 0.22; Supplementary figure S3). One Glx study (Sanacora et al., 2004) was identified as an outlier and was excluded from the analysis (before exclusion *I*^2^ = 71.6%, *p* < 0.001; after exclusion, *I*^2^ = 36.6%, *p* = 0.15; Supplementary figure S3).

**Table 2:** Meta-analysis studies. Each study included in the GABA and Glx meta-analyses. Magnetic field strength in Tesla, MRS sequence, which transmitters were measured, and the referencing approach used are given for each.

| Study | Field (T) | Sequence | Neurotransmitter | Reference |
| --- | --- | --- | --- | --- |
| Sanacora et al. (1999) | 2.1 | J-editing | GABA | Creatine |
| Kugaya et al. (2003) | 2.1 | J-editing | GABA | Creatine |
| Mirza et al. (2004) | 1.5 | PRESS | Glx | Phantom |
| Sanacora et al. (2004) | 2.1 | J-editing | GABA & Glx | Creatine |
| Rosenberg et al. (2005) | 1.5 | PRESS | Glutamate | Creatine |
| Bhagwagar et al. (2008) | 3 | MEGA-PRESS | GABA & Glx | Creatine |
| Price et al. (2009) | 3 | PRESS | GABA | Water |
| Abdallah et al. (2014) | 4 | J-editing | GABA & Glutamate | Creatine |
| Shaw et al. (2013) | 3 | MEGA-PRESS | GABA | Water |
| Godlewska et al. (2014) | 3 | SPECIAL | GABA & Glutamate | Creatine +<br>phosphate |
| Godlewska et al. (2018) | 7 | Semi-LASER | Glutamate | Water |
| This study (2020) | 3 | MEGA-PRESS | GABA & Glx | Water |

A comparison of occipital GABA concentration differences between MDD patients and healthy control subjects form seven studies was performed. Based on a random-effects model meta-analysis, there was no significant difference in occipital GABA concentration between MDD and control groups (SMD = −0.24, permutation test 95% CI = −0.56 to 0.08, *p* = 0.11; Figure 4A). Visual inspection of the funnel plot suggested (Figure 4B) that there was a minimal risk of publication bias as many studies reported non-significant results. A meta-regression of the meta-analysis was conducted to test the effect of publication year and magnetic field strength on reported occipital GABA concentration differences. This showed that publication year (*F*_(1,6)_ = 22.22, *p* = 0.003; Figure 4C), but not magnetic field strength (*F*_(1,6)_ = 5.49, *p* = 0.058; Figure 4D), was positively related with reported group differences for occipital GABA.

**Figure 4:**
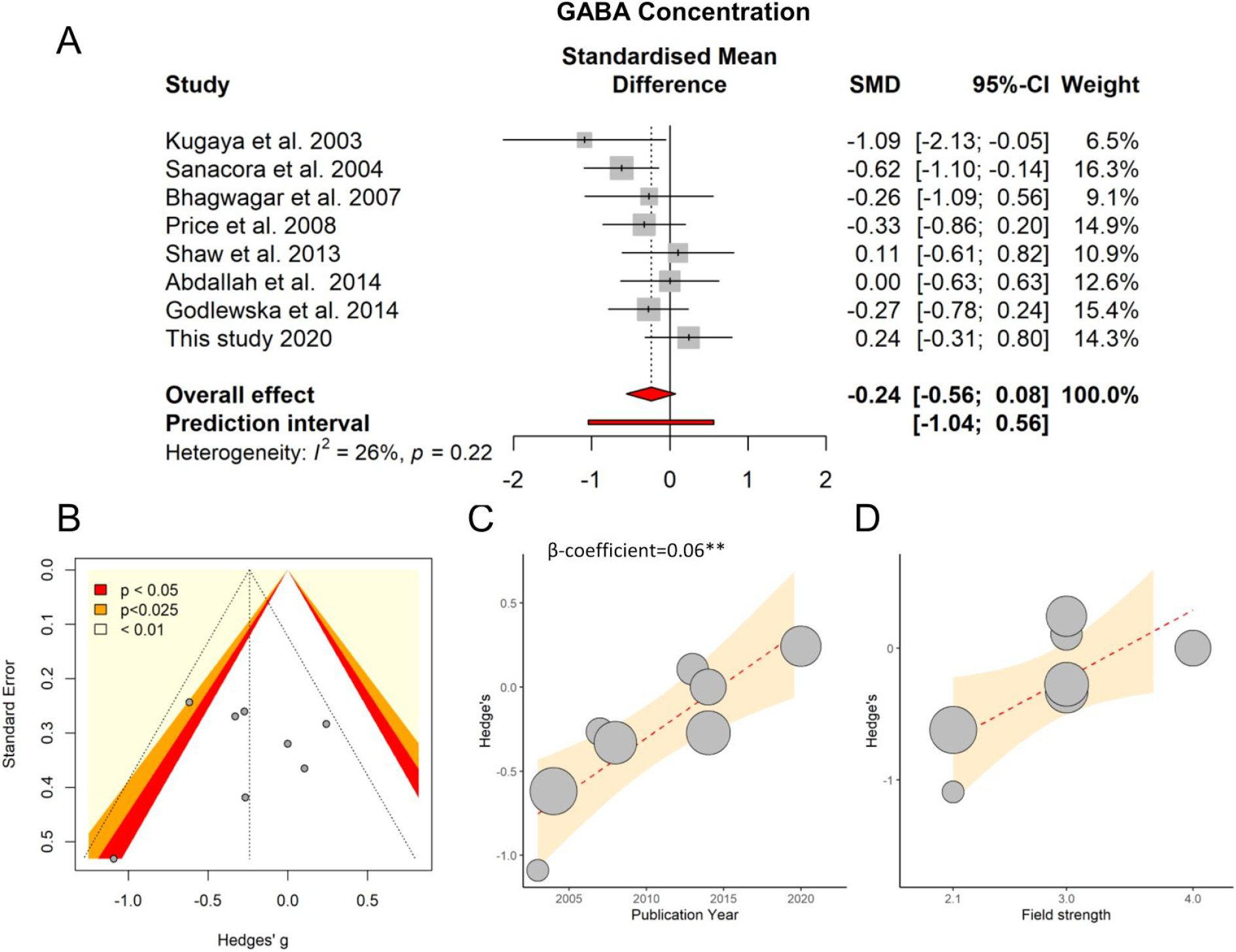
Meta-analysis of occipital GABA differences. (A) Forest plot of effect sizes from each study. The overall effect is indicated in red. (B) Funnel plot indicating a minimal risk of publication bias. (C) Regression of group difference effect size and year of publication. (D) Regression of group difference effect size and magnetic field strength. Best fit straight lines are shown with their CI. Bubbles represent the weight assigned to each study. ** denotes *p* < 0.001.

The same approach was then applied to reported Glx differences from seven studies. No significant difference in occipital Glx concentrations between MDD and control groups was found (SMD = −0.28, permutation test 95% CI = −0.70 to 0.13, *p* = 0.11; Figure 5A). A potential publication bias may exist based on the presence of studies with small sample sizes and non-significant results (Figure 5B). In contrast to GABA, there was no evidence that publication year (*F*_(1,5)_ = 0.84, *p* = 0.40; Figure 5C) influenced reported Glx group differences, nor was there any relationship with magnetic field strength (*F*_(1,5)_ = 0.20, *p* = 0.67; Figure 5D).

**Figure 5:**
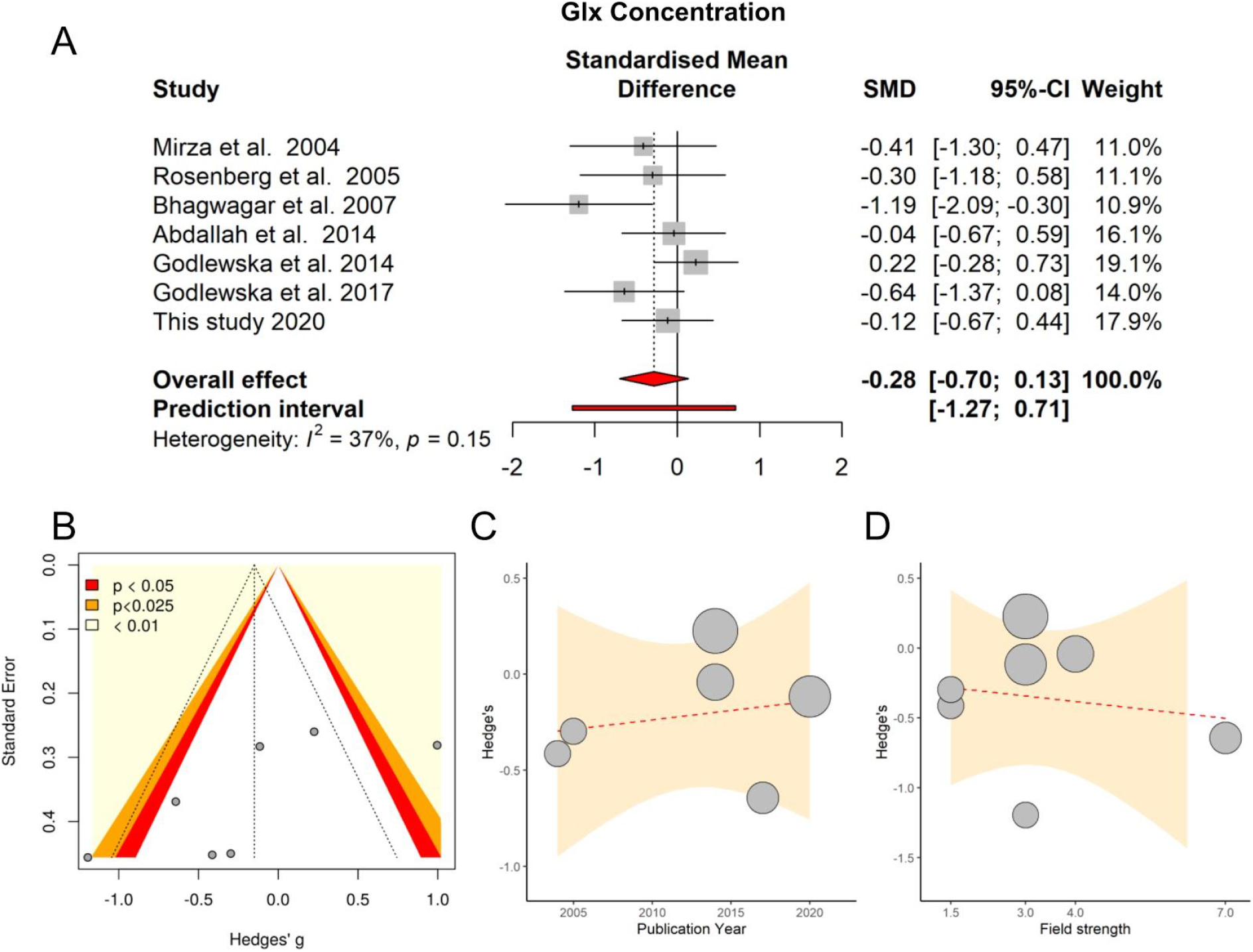
Meta-analysis of occipital Glx differences. (A) Forest plot of effect sizes from each study. The overall effect is indicated in red. (B) Funnel plot indicating a minimal risk of publication bias. (C) Regression of group difference effect size and year of publication. (D) Regression of group difference effect size and magnetic field strength. Best fit straight lines are shown with their CI. Bubbles represent the weight assigned to each study.

## 4. Discussion

The observed lack of difference in occipital GABA contrasts with early work highlighting substantial reductions in MDD (Kugaya et al., 2003; Sanacora et al., 2004, 1999) but coheres with more recent studies suggesting no difference between patients and controls (Bhagwagar et al., 2004; Godlewska et al., 2014; Price et al., 2009; Shaw et al., 2013). This includes a recent study that included patients with bipolar depression that was excluded from the meta-analysis (Knudsen et al., 2019). Our meta-analysis result demonstrating an influence of publication year on reported group differences suggests that the early studies may have produced false-positives due to methodological issues that are not seen in later studies. This highlights the need to replicate findings within the psychiatric MRS literature as scanning technology and analysis techniques improve.

GABA concentrations not being altered in the occipital cortex raises interesting questions regarding mechanisms for alterations in that system in MDD given the evidence that there are alterations in other brain regions. A recent meta-analysis taking the brain as a whole showed reductions in GABA concentrations in patients (Godfrey et al., 2018). The same work also showed a reduction in the anterior cingulate cortex when integrating results from that region specifically. Other individual MRS studies have also suggested changes in the dorsolateral and dorsomedial prefrontal cortices (Bhagwagar et al., 2008; Hasler et al., 2007). These regions lie within the salience and executive control networks, which are known to be functionally altered in MDD (Northoff, 2016), pointing to potential network-specific changes in GABA concentrations. Although the mechanisms behind such changes remain to be identified, there is increasing evidence for localised interactions between stress and GABAergic interneuron function leading to depressive symptoms (Fogaça and Duman, 2019; Girgenti et al., 2019).

Both our MRS analysis and meta-analysis also revealed no difference in occipital Glx concentrations in MDD. No change was observed in GABA/Glx ratios either. Prior work looking at occipital Glx in MDD has given varied results, with some studies finding increases (Sanacora et al., 2004), some decreases (Bhagwagar et al., 2008), and others no difference. Our negative result from the occipital cortex fits with recent work using high-field strength MRS to look at glutamate in other brain regions that also found no difference between MDD patients and controls (Godlewska et al., 2018). It may be noted, however, that the glutamate-glutamine system plays a metabolic as well as neurotransmission role and that there may be changes to neuronal energy production that are not visible to the MRS methods employed (Abdallah et al., 2014) and which may be relevant to the aetiology of depression (Allen et al., 2018; Morava and Kozicz, 2013).

The specific locations of excitatory and inhibitory neurotransmitter changes in depression may be informative from the point of view of active inference theories of depression (Kube et al., 2020). According to these, the processes whereby signals representing sensory inputs are compared to the brain’s internal model become distorted. This could in principle occur at different stages of the process: in the encoding of sensory signals; in the properties of the internal model; in the generation of prediction errors; or in the model updates and behaviour selection resulting from them. The apparent lack of MRS-based evidence for visual cortex changes suggest that sensory signals themselves are not affected. The previously described changes in the anterior cingulate and prefrontal cortices (Godfrey et al., 2018) would instead point to issues with the internal model or prediction error generation, processes that are thought to be associated with these regions (Alexander and Brown, 2019; Pezzulo et al., 2014).

The current work has a number of limitations. Firstly, the patients included in the MRS study were predominantly female and so we also cannot rule out sex effects on occipial metabolites. This could include hormonal influences on GABA concentrations (Epperson et al., 2002), although it would be unlikely that this effect would be consistent enough across all participants to drive the results. Secondly, the number of studies included in the meta-analysis is lower than optimum for reliability and methodological heterogeneity between studies is inherent (voxel size, MRS sequence, etc). Finally, although the consistent MRS and meta-analysis results provide strong evidence for there being no change in occipital GABA when taking MDD patients as a whole, increasing evidence points to different disease subtypes being subsumed within the MDD category (Drysdale et al., 2017; Woelfer et al., 2019). These subtypes may have different biological underpinnings meaning that occipital neurotransmitter changes could be present in some but not others (Beijers et al., 2019). This would be consistent with the finding of no effect across all participants as a whole.

In conclusion, MRS data from MDD patients and controls were compared to identify changes in inhibitory and excitatory neurotransmitter concentrations in the occipital cortex. This analysis was then complemented by a meta-analysis of similar prior studies. Both analyses gave consistent evidence that occipital GABA+ and Glx concentrations do not differ between patients with MDD and controls. This provides a synthesis of prior work and clarifies inconsistencies in the literature, suggesting that early reports of occipital GABAergic changes in MDD are likely to not have been correct.

## Data Availability

The data used in this work are freely available

https://osf.io/3znd9/

## Acknowledgments

The authors would like to thank all participants for their time and effort. This work was supported by funding from the Taiwan Ministry of Science and Technology to TJL (104-2420-H-038-001-MY3; 105-2632-H-038-001-MY3); TYH (106-2410-H-038-004-MY2), and NWD (107-2410-H-038-004-MY2; 108-2410-H-038-008-MY2).

## Author contributions

NWD, TYH, & TJL conceived of the study; NWD & VT planned the analysis; PZC & HCL collected MRS data; VT collated the meta-analysis data; VT conducted the analysis; VT & NWD wrote the manuscript; all authors reviewed the manuscript prior to submission. The authors declare no conflicts of interest.

## Data availability

The data used in this work are available at https://osf.io/3znd9/.

## Supplementary materials

**Table S1:** Patient treatment information. Time since first onset (in years), average duration of current episode (in months), and medications prescribed to each MDD patient. Missing data is marked as N/A. Abbreviations: SNRI = serotonin-norepinephrine reuptake inhibitor; Non-BDZ = non-benzodiazepine hypnotic; ATA = atypical antipsychotic; MRA = melatonin receptor agonist; BDZ = benzodiazepine; SSRI = selective serotonin reuptake inhibitor; NDRI = norepinephrine-dopamine reuptake inhibitor; SARI = serotonin antagonist and reuptake inhibitor

| Patient ID | Time since first onset (years) | Duration of current episode (months) | Medications |
| --- | --- | --- | --- |
| p01 | 4 | 1.5 | SNRI, Non-BDZ, BDZ, ATA |
| p02 | N/A | 6 | NDRI |
| p03 | 9 | 2 | MRA, non-BDZ |
| p04 | 2 | 6 | SSRI, BDZ |
| p05 | 36 | 2 | BDZ, non-BDZ, NDRI |
| p07 | 3 | 4 | ATA, SSRI |
| p08 | 8 | 1 | NDRI |
| p09 | 6 | 23 | MRA |
| p10 | 12 | 3 | NDRI |
| p11 | 10 | 6 | SSRI, ATA |
| p12 | 3 | 3 | SSRI, ATA, NDRI |
| p13 | 4 | 1 | SNRI, ATA |
| p14 | 0 | 3 | SSRI |
| p15 | 33 | 3 | SNRI, non-BDZ, SARI |
| p18 | 1 | 24 | SSRI |
| p21 | 1 | 8 | SNRI, non-BDZ, SARI, ATA |
| p23 | 9 | 6 | SSRI, BDZ |
| p25 | 1 | 4 | SSRI |
| p26 | 20 | 3 | SSRI, BDZ |
| p27 | 25 | N/A | SSRI |
| p28 | 10 | 88 | SNRI, ATA, BDZ, SARI |
| p29 | 9 | 3 | SSRI, BDZ, ATA |
| p30 | 27 | 2 | non-BDZ, BDZ |
| p31 | 20 | 13 | NDRI |
| p35 | 14 | 18 | SNRI, BDZ, non-BDZ, ATA |
| p37 | 6 | 3 | SNRI, ATA |
| p38 | 38 | N/A | Treatment naive |
| p39 | 5 | 4 | NDRI, BDZ |
| p40 | N/A | 3 | NDRI, ATA |
| p42 | 4 | 18 | SNRI, BDZ, ATA |
| p44 | 0 | 2 | non-BDZ, BDZ, ATA |

**Figure S1:**
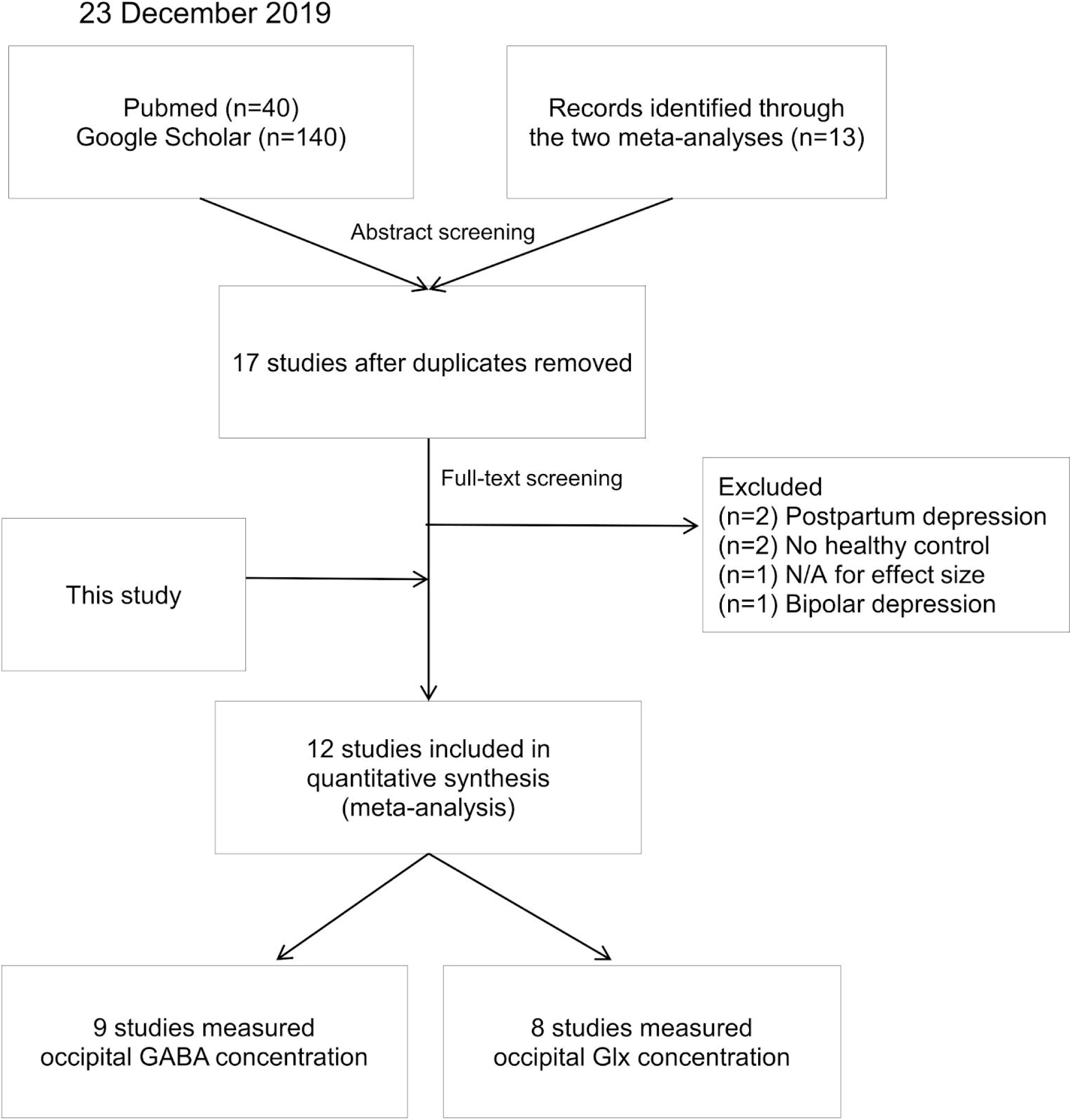
Study selection process. Search results were obtained on 23rd December 2019 from Pubmed, Google Scholar and records identified through the two previous meta analyses. These were screened using their abstracts. After removing all duplicates, 17 selected studies underwent another screening process using their full text. A total of nine studies for occipital GABA concentration and eight studies for occipital Glx concentrations were included in the two meta-analyses.

**Table S2:** Excluded studies. Studies excluded after full text screening.

| Study | Reason for exclusion |
| --- | --- |
| Deligiannidis et al. (2019) | Peripartum depression |
| Epperson et al. (2006) | Peripartum depression |
| Knudsen et al. (2019) | Includes bipolar depression |
| Murrough et al. (2010) | Insufficient information for effect size |
| Sanacora et al. (2002) | No healthy control |
| Sanacora et al. (2003) | No healthy control |

**Figure S2:**
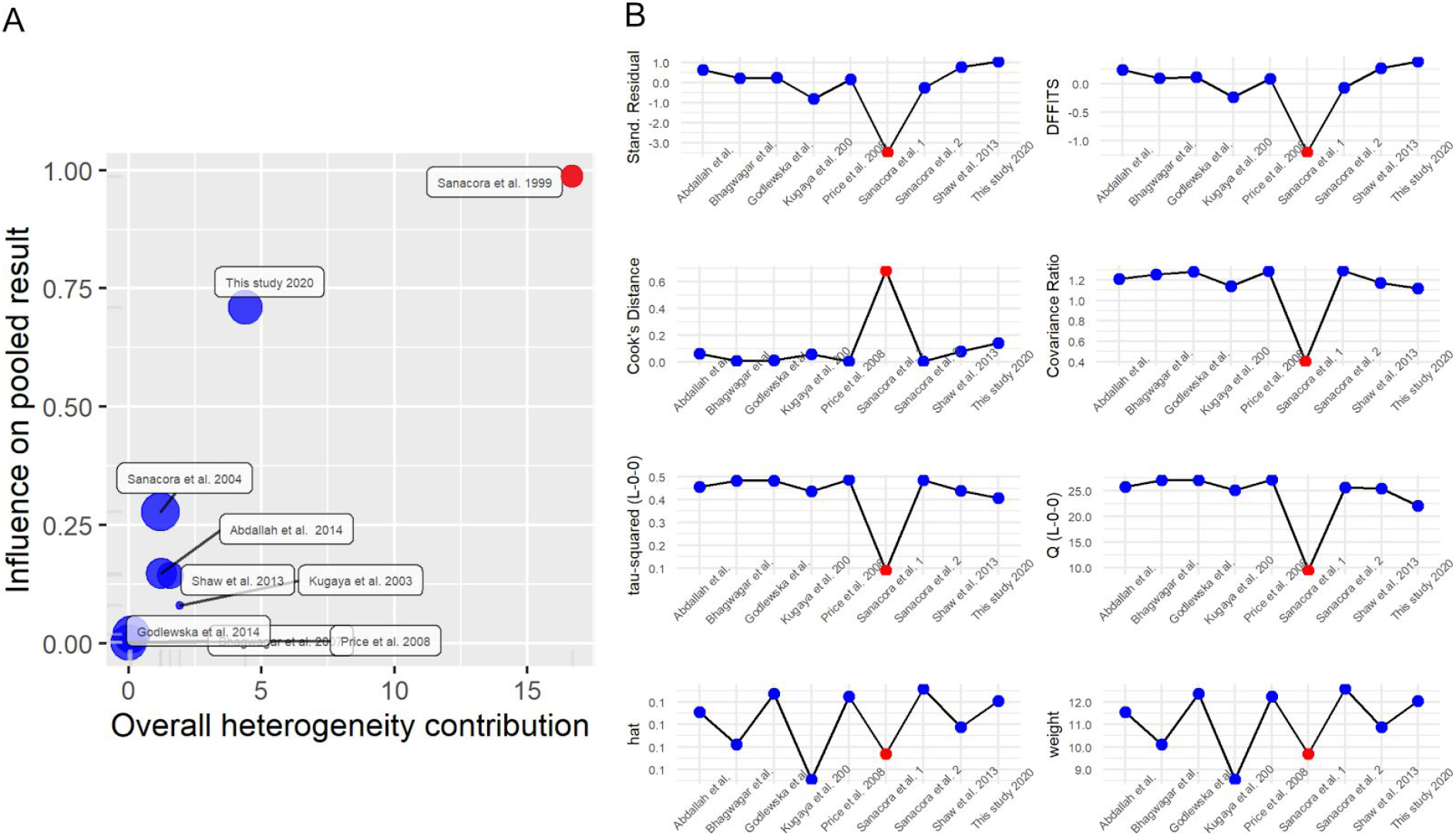
Influence diagnostics for occipital GABA concentration studies. (A) Illustration of the overall heterogeneity contribution on influence on pooled effect size of each study. (B) Plots of (1) externally standardized residuals, (2) DFFITS values, (3) Cook’s distances, (4) covariance ratios, (5) leave-one-out values of the test statistics for heterogeneity, (7) hat values, and (8) weights of the studies. One study (shown in red) was identified as an outlier.

**Figure S3:**
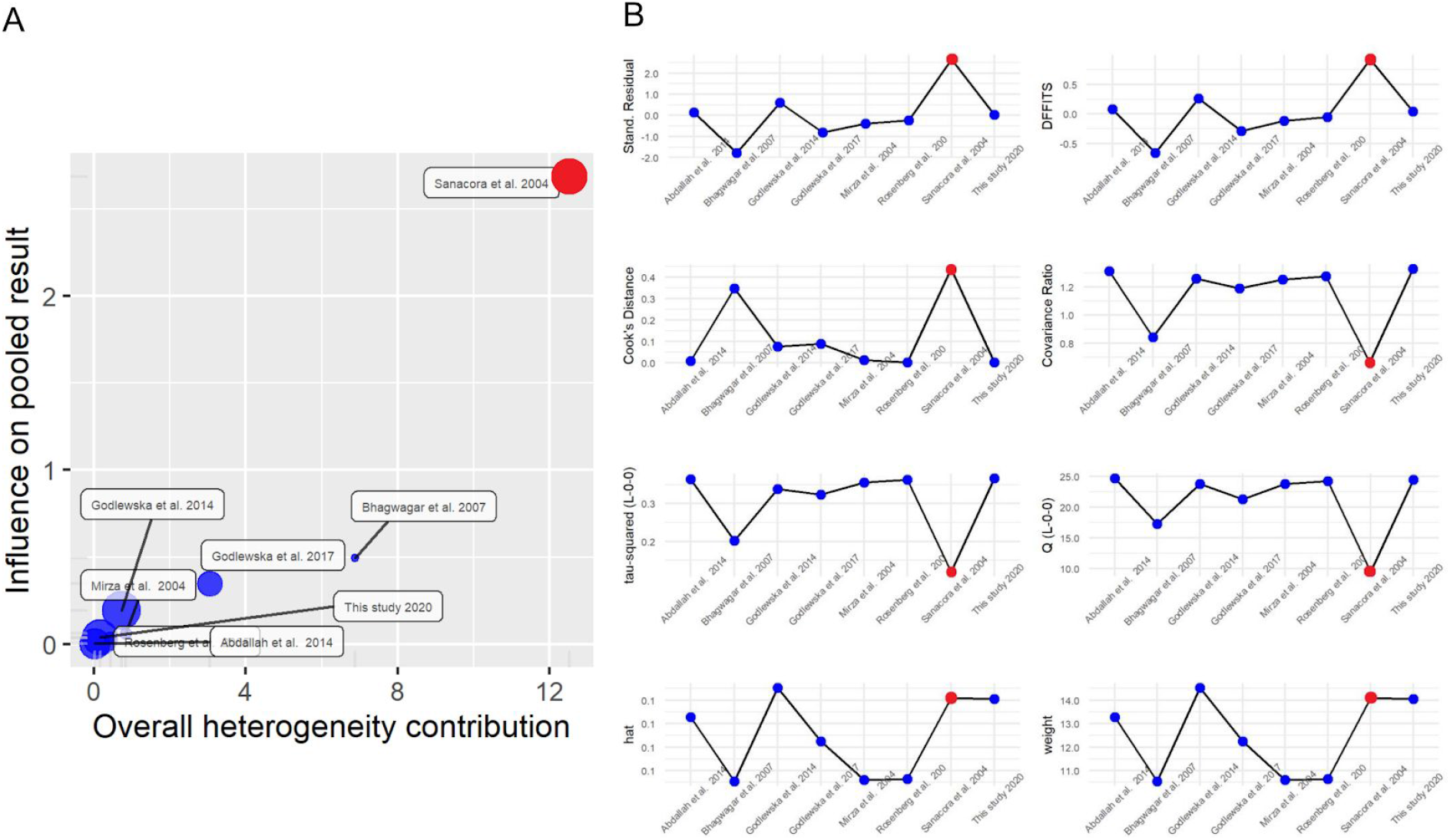
Influence diagnostics for occipital Glx concentration studies. (A) Illustration of the overall heterogeneity contribution on influence on pooled effect size of each study. (B) Plots of (1) externally standardized residuals, (2) DFFITS values, (3) Cook’s distances, (4) covariance ratios, (5) leave-one-out values of the test statistics for heterogeneity, (7) hat values, and (8) weights of the studies. One study (shown in red) was identified as an outlier

**Figure S4:**
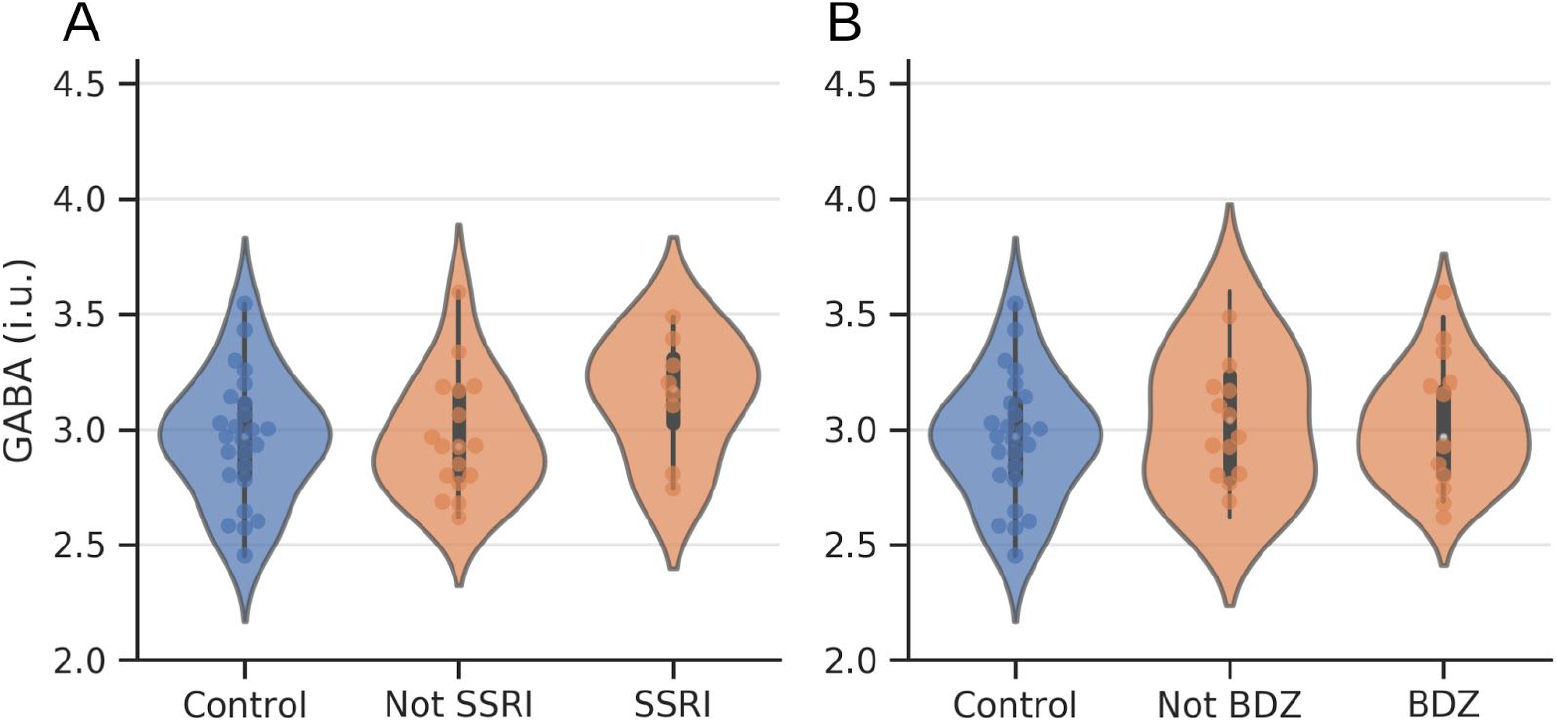
Medication effects on occipital GABA+. (A) GABA+ estimates for controls (n = 25), patients not using SSRIs (n = 17), and patients using SSRIs (n = 8). (B) GABA+ estimates for controls (n = 25), patients not using BDZs (n =12), and patients using BDZs (n = 13). No differences were seen between groups. MDD patients are shown in orange and controls in blue. Box plots indicate medians and interquartile range. BDZ = benzodiazepine; SSRI = selective serotonin reuptake inhibitor.

